# Scabies outbreak management in refugee/migrant camps across Europe 2014-17: a retrospective qualitative interview study of healthcare staff experiences and perspectives

**DOI:** 10.1101/2021.04.28.21256211

**Authors:** N. A. Richardson, J. A. Cassell, M. G. Head, S. Lanza, C. Schaefer, S.L. Walker, J. Middleton

## Abstract

**Background:** Scabies outbreaks were common in formal and informal refugee/migrant camps across Europe in 2014-17. This qualitative study aimed to provide insight into the experiences and perspectives of healthcare staff who treated scabies or managed outbreaks in these camps.

**Methods:** Recruitment was primarily through online networks of healthcare staff involved in medical care in refugee/migrant settings. Retrospective semi-structured telephone interviews were conducted, transcribed, and qualitative framework analysis carried out.

**Results:** Twelve participants who had worked in camps across seven European countries were interviewed. They reported that in the camps they had worked scabies diagnosis was primarily clinical, and without dermatoscopy, and treatment and outbreak management varied highly. Seven participants stated scabicide treatment was provided in camps whilst they were there, the remaining five reported only symptomatic management was offered. They described the camps as difficult places to work, with poor standards of living experienced by residents. Key perceived barriers to scabies control were (i) lack of Water, Sanitation and Hygiene facilities, specifically: absent/ limited showers (difficult to wash off irritant topical scabicides); inability to wash clothes and bedding (may have increased transmission/re-infestation), (ii) social factors: language; stigma; treatment non-compliance; mobility (interfering with contact tracing and follow-up treatments), (iii) healthcare factors: scabicide shortages and diversity; lack of examination privacy; staff inexperience, (iv) organisational factors: overcrowding; ineffective inter-organisational coordination; lack of support and maltreatment by state authorities (e.g. not providing basic facilities, and obstruction of self-care by camp residents and of aid efforts by NGOs).

**Conclusions:** We recommend development of accessible scabies guidelines for camps, use of consensus diagnostic criteria, and oral ivermectin mass treatments. In addition, as much of the work described was by small, volunteer staffed NGOs, we should reflect how we in the wider healthcare community can better support such initiatives, and those they serve.

## 1.0 INTRODUCTION

Scabies is a stigmatised contagious skin condition caused by infestation with the mite *Sarcoptes scabiei* [1]. Transmission is mainly skin to skin, less commonly via fomites such as bedding [2]. Symptoms begin 3-6 weeks after first infestation, but as early as one day after re-infestation [3]. Secondary bacterial infections (with *Staphylococcus aureus* and/or group A *Streptococcus*) are common [4,5], with potential for serious long-term health impacts, including chronic rheumatic heart disease and chronic kidney disease [6-8]. Diagnosis is normally by clinical examination, sometimes with the aid of dermatoscopy [3]. Definitive diagnosis has been classified as requiring ‘clinical features suggestive of scabies… plus a visible mite under dermatoscopy … or a skin scraping or biopsy with identified mites, mite eggs, or faeces’ [9]. Simultaneous treatment for diagnosed individuals and all close contacts is required [10]. This normally consists of either topical scabicides applied to the entire body or oral ivermectin, in both cases often given twice (one week apart) [11]. Advice on environmental decontamination varies [11]. There are an estimated 455 million annual cases of scabies, which is designated by WHO as a Neglected Tropical Disease [12]. The highest burden is in the tropics but in low-burden regions (such as Europe) scabies remains common and emerges as a public health problem when outbreaks occur in institutions [9,12].

Conflicts in Africa and the Middle East triggered increased numbers of people to migrate to Europe in recent years [13-15]. By the end of 2016 European countries hosted 5.2 million refugees, 2.9 million (largely from Syria) in Turkey alone [13]. As a result, during the period 2014-17 many formal refugee/migrant camps and reception centres, and informal (often illegally occupied) camps came into being or markedly expanded (Figure 1). Camp conditions varied, but an environmental health assessment of an informal one in Calais found that, (1) residents had to queue to access the 400 daily shower slots for the population of c.3000 people, which functioned only 3 h daily; (2) healthcare services were limited to clinics operating from caravans or makeshift structures, largely staffed by volunteers [16]. Formal reception centres have been described as having better water and sanitation facilities; with daily cleaning and some providing clothing on arrival. However, they can still become overcrowded by unexpected large influxes. Lack of sustainable funding can cause staffing and medicine shortages, resulting in ineffective care [14].

**Figure 1.**
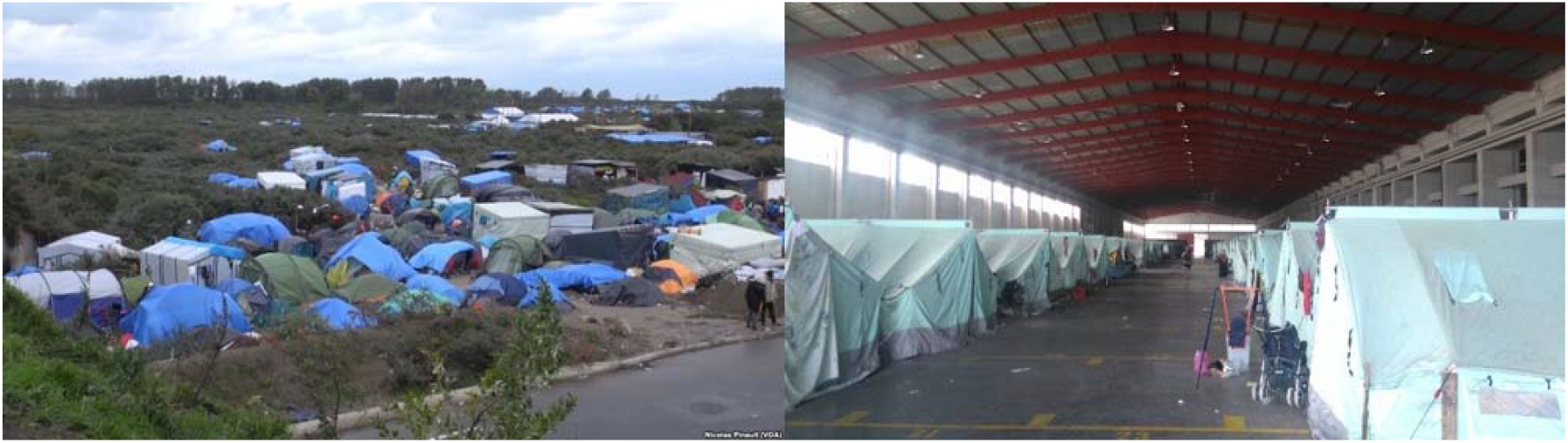
Informal and formal refugee/migrant camps Left: informal camp in Calais, France, 2015 (VOA News - Nicolas Pinault (2015), public domain, original: https://web.archive.org/web/20210406154625/ https://commons.wikimedia.org/wiki/File:Calais2015a.jpg). Right: formal camp Thessaloniki, Greece, 2016 (Konstantinos Stampoulis (2016), used under CC BY-SA 4.0 which also applies to this figure, original: https://web.archive.org/web/20210406155208/ https://commons.wikimedia.org/wiki/File:Refugee_hotspot_Fessas_(3).jpg).

Scabies was one of the most frequently reported medical conditions in these settings [16-19]. In Germany, it was the third most common outbreak type (behind chickenpox and measles) in shelters for asylum seekers [19]. In France, Doctors of the World UK estimated in 2015 that up to 40% of those seeking their care in the Calais ‘Jungle’ camp had scabies (Cooper C. 2015, Appendix, p.2). Scabies outbreaks in reception centres and camps have been widely reported in news and social media, and a media analysis by Seebach *et al*. [20] described much of this coverage as framed with an anti-migrant narrative that stigmatises those affected by scabies whilst positing migrants/refugees as an invasive health threat to European countries. Multiple camps in France have been evicted with the public justification of responding to scabies outbreaks (examples of newspaper coverage: Lichfield J 2014; Newton J 2015, Appendix, p. 2). Despite all this, to our knowledge no study prior to ours has been published explicitly on scabies outbreak diagnosis, treatment, and management in refugee/migrant camps in Europe during this period. Further, we are unaware of any published study on healthcare staff perspectives on barriers and facilitators to treating and managing scabies in refugee camps globally.

This qualitative study aimed to provide an insight into the experiences and perspectives of healthcare staff who have treated scabies or managed associated outbreaks in refugee/migrant camps in Europe 2014-17. Specifically, to describe (i) methods used to diagnose, treat, and manage scabies, and (ii) camp characteristics and perceived barriers and facilitators to effective scabies outbreak management in these settings.

## 2.0 METHODS

A retrospective qualitative study was carried out using semi-structured telephone interviews.

### 2.1 Participant selection and recruitment

Healthcare staff from state and non-governmental organisations who had treated scabies in refugee/migrant camps in Europe, and/or managed associated outbreaks in the previous three years were eligible. As far as possible within time and resource constraints, we interviewed a diverse sample by age, sex, organisation, and location. We recruited by contacting:

i. relevant healthcare staff known to the research team;
ii. individuals publishing research on medical care in refugee/migrant settings;
iii. through online networks of healthcare staff involved in medical care in refugee/migrant settings, including the alumni network of the European Centre for Disease Control fellowship program (field epidemiology path) and groups hosted on facebook.com. To target the latter a public page (https://web.archive.org/web/20180523163917/https://www.facebook.com/scabies researchproject/) was shared to ten relevant facebook.com hosted groups (Table S1, Appendix, p.2) and messages sent to members whose posts indicated they fitted the inclusion criteria.

An information sheet was provided to potential participants, followed with a verbal explanation prior to interviews. Explicit verbal consent was given over the telephone, audio-recorded and stored separately from the interviews.

### 2.2 Data Collection, Processing, and Analysis

Prior to interviews, participants gave data on personal characteristics and experience. Audio-recorded, semi-structured telephone interviews were conducted from a private room in the Department of Primary Care and Public Health at Brighton and Sussex Medical School by first author NR November 2017 to February 2018. Interviews followed a topic guide based on published literature and guidance from experts in scabies (epidemiology, JAC; dermatology, SLW; medical acarology, JM), and were scheduled to be around 40mins long. However, so as to provide sufficient information power [21] participants were told they could speak longer as they felt necessary, and to not feel precluded by the pre-written topic guide from relating any of their experiences and perspectives they though relevant to the study aims. Data collection and thematic analysis (by NR) were conducted simultaneously so as to determine when data saturation was reached [22,23], which was prospectively defined as being no new themes emerging in three consecutive interviews. Once data saturation was observed no new interviews were conducted as the research team could ‘be reasonably assured that further data collection would yield similar results and serve to confirm emerging themes and conclusions’ [24]. Transcription was undertaken by professional service (9 of 12 interviews), or by NR (3 of 12). Anonymized recordings and transcriptions were stored on secure password-protected university network storage (retained for three years after study completion). NR coded text in NVivo 11 (QSR International, Melbourne), generated an initial framework matrix and exported it to Microsoft Excel, in which qualitative framework analysis was completed (as outlined by Gale *et al*. [25]). Groups of meaningful concepts were sought in the data and arranged hierarchically into main themes and subthemes. Each transcript was studied again twice for more evidence of the concepts. Subsequently author JM reviewed the reported themes and subthemes and read the transcripts, finding no new relevant themes or subthemes than those grouped by NR and thus reaching consensus.

Brighton and Sussex Medical School Research Governance and Ethics Committee approved the research (ER/BSMS3398/1). We have reported our work in line with the Standards for Reporting Qualitative Research guidelines [26] (researcher characteristics and checklist, Appendix, p.2-3).

## 3.0 RESULTS

Recruitment halted after 12 interviews as data saturation was reached (no new themes emerged in the last three interviews). Interviews lasted 34-71min (mean 47).

### 3.1 Study Population

12 participants were recruited with relevant experience in 2014-17: four doctors, four nurses, three allied health workers (AHWs: an ECG technician, podiatrist, and a first aider), and one medical student. Six had worked in informal settings, nine in formal camps, within seven European countries: Greece, France, the Netherlands, Serbia, Belgium, Turkey, Macedonia (Figure 2). Ten were NGO volunteers, one an independent volunteer, one was employed by a public health service. Experience in the camps ranged from 4 d to 2.5 y (median 84 d, IQR 233-251 d) (participant characteristics detailed in Table S3, Appendix, p.4).

**Figure 2.**
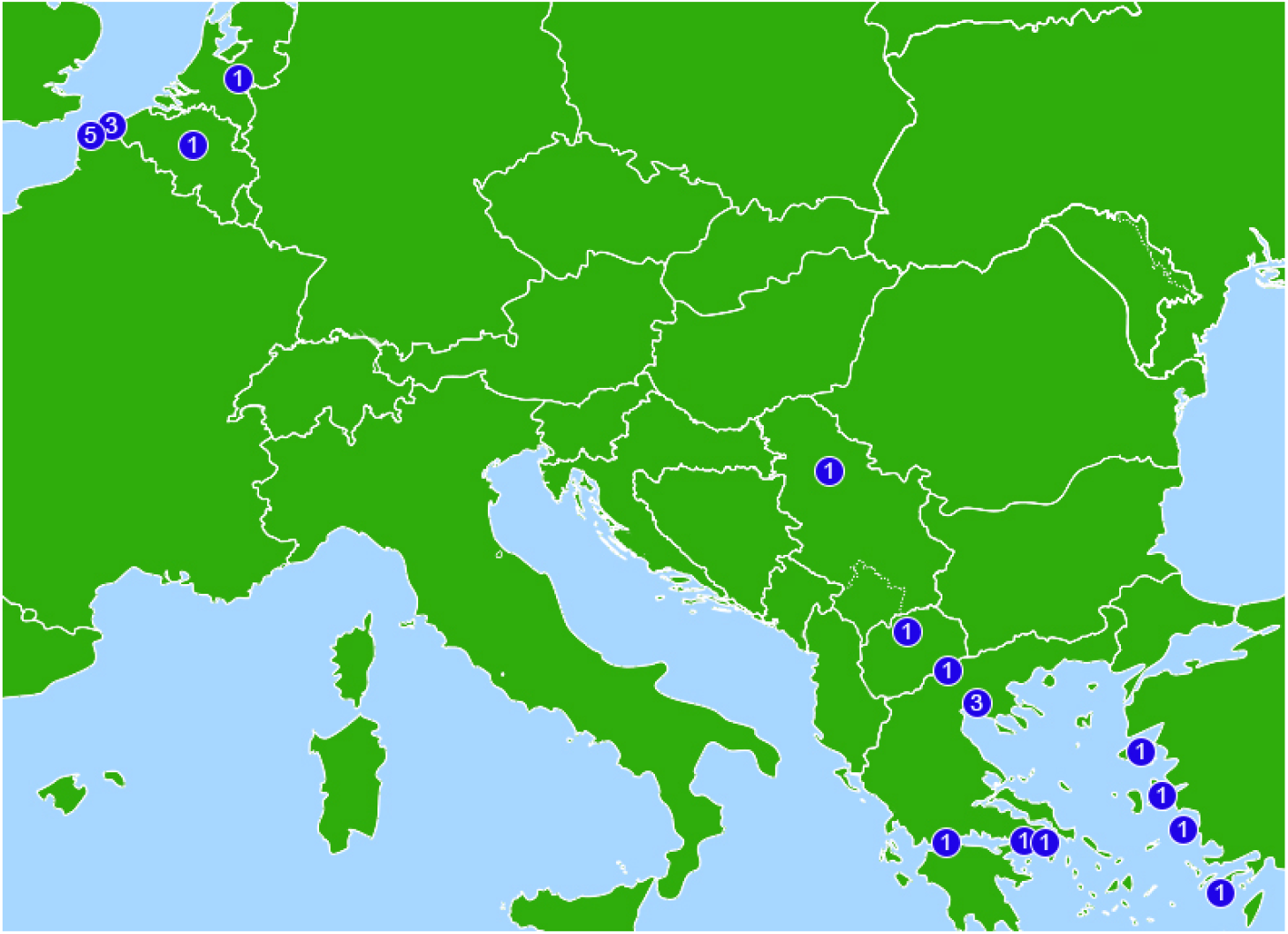
Camp locations and number of participants (n=12) with experience at each site Distribution of camps follows eastern entry migration routes into Europe, and clustering at the English Channel by those seeking to enter the UK. Left to right: Calais, France, 5 participants all active at informal (IF) camps; Dunkirk, France, 3 all IF; Brussels, Belgium, 1 formal (F); Heumensoord, Netherlands, 1 F; Belgrade, Serbia, 1 IF; Macedonia, 1 IF; Patras, Greece, 1 IF; Idomeni, Greece, 1 IF; Thessaloniki, Greece, 3 – 2 F, 2 IF; Athens, Greece, 1 IF; Attica, Greece, 1 F; Lesbos, Greece, 3 –1 F, 2 IF; Çesme, Turkey, 1 IF; Samos, Greece, 2 F; Tilos, Greece, 1 F. (Figure uses adapted basemap by Julio Reis (2006) created using public domain mapping from the European Environment Agency, use and changes made under CC BY-SA 3.0 which also applies to this image. Original basemap: https://web.archive.org/web/20161019140631/ https://commons.wikimedia.org/wiki/File:Europe_biogeography_blank.svg.)

### 3.2 Diagnosis, treatment, and outbreak management

Scabies diagnosis and treatment are summarized in Table 1, and outbreak management in Table 2.

**Table 1.**
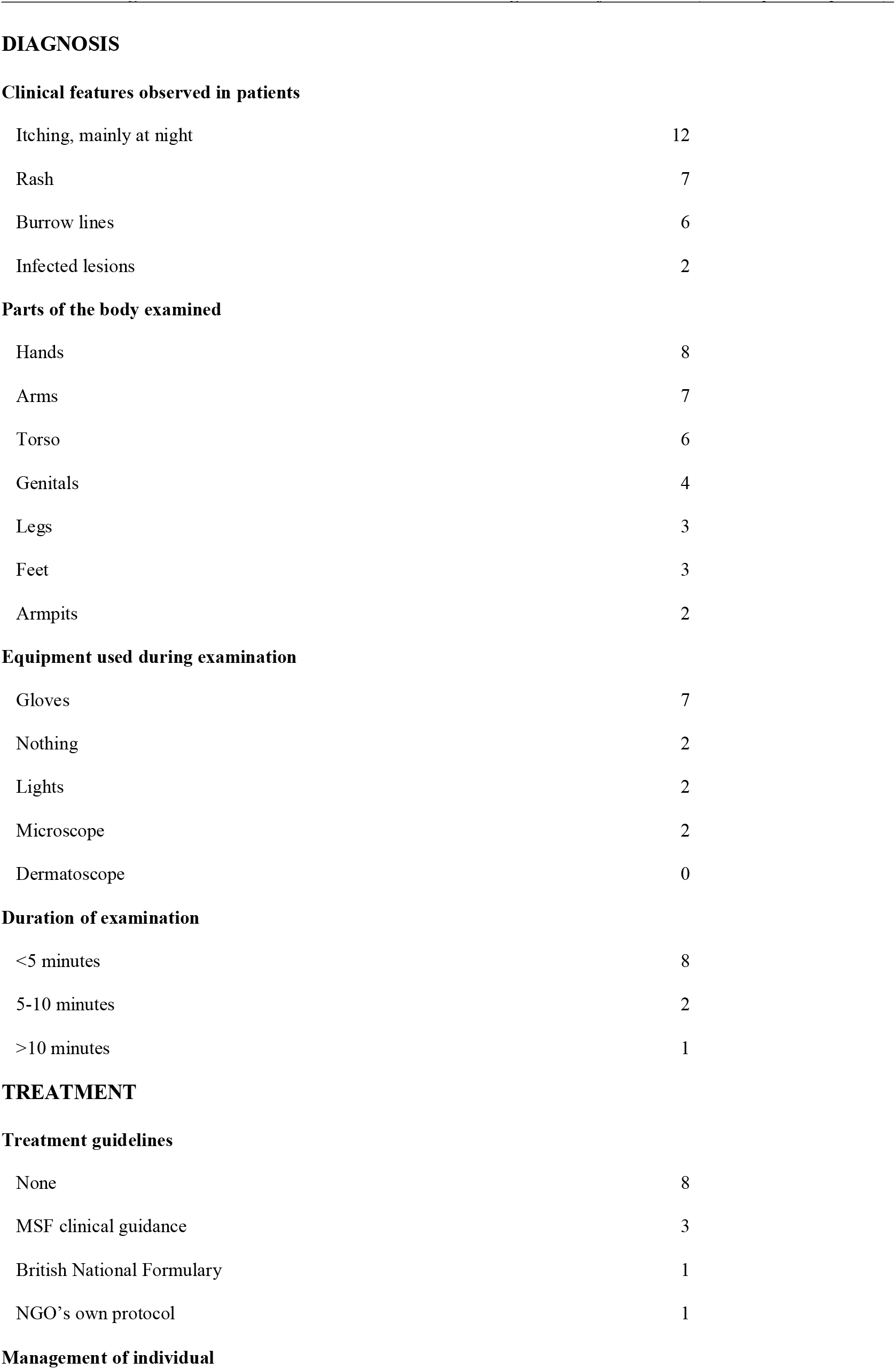
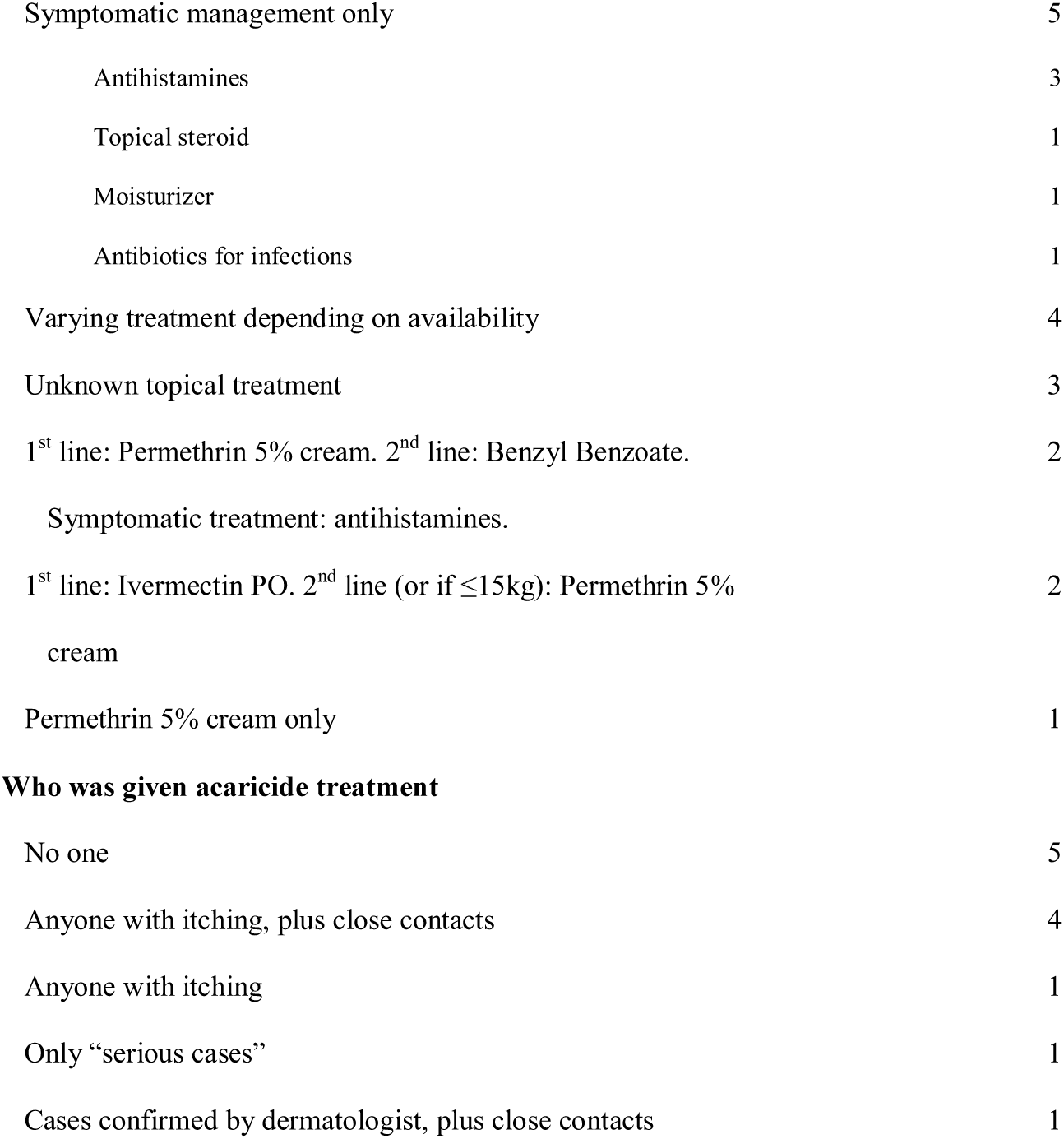
Diagnosis, treatment, and outbreak management of scabies (n= 12 participants)

**Table 2.**
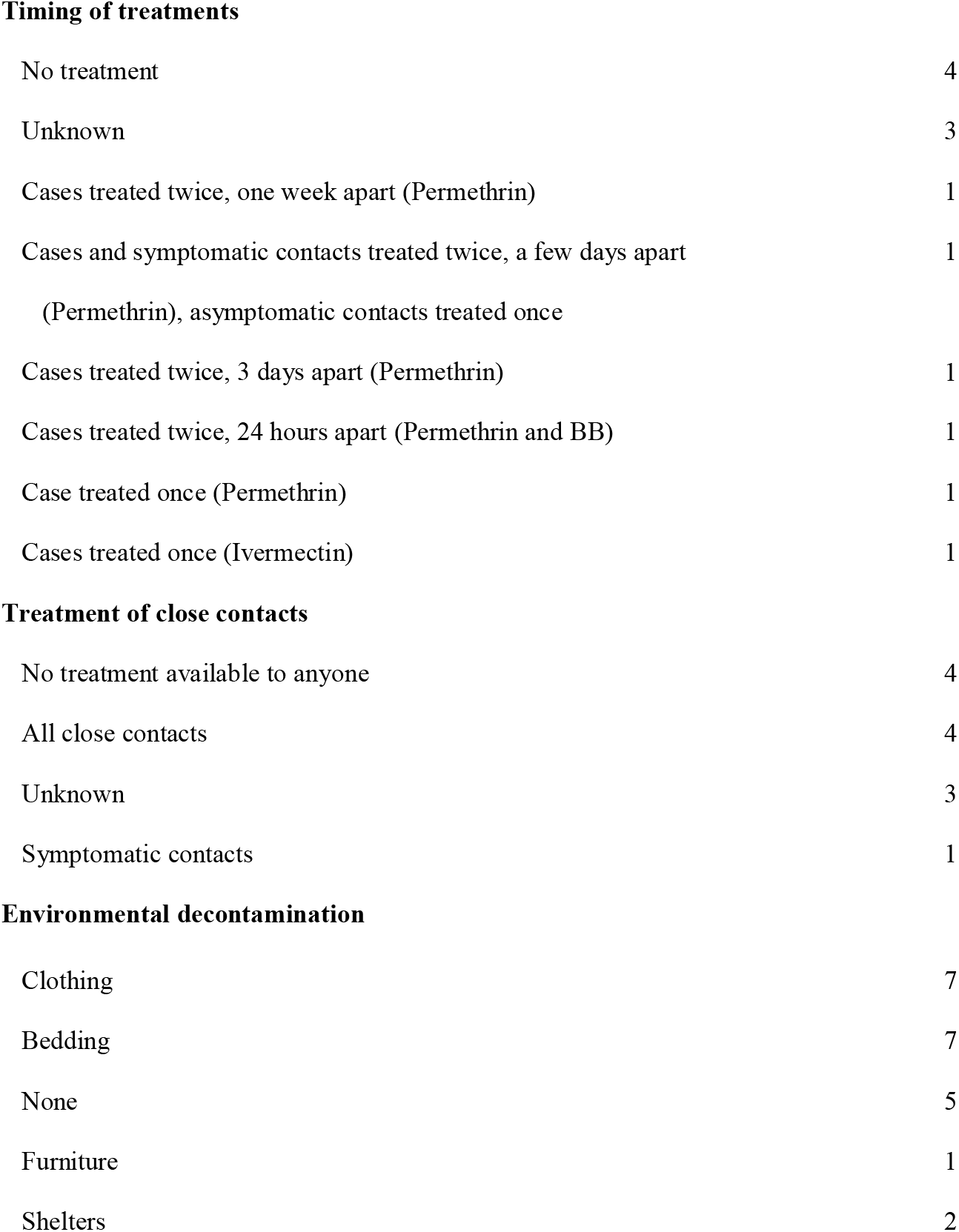
Outbreak management of scabies (n= 12 participants)

|  |  |
| --- | --- |
| No treatment | 4 |
| Unknown | 3 |
| Cases treated twice, one week apart (Permethrin) | 1 |
| Cases and symptomatic contacts treated twice, a few days apart<br>(Permethrin), asymptomatic contacts treated once | 1 |
| Cases treated twice, 3 days apart (Permethrin) | 1 |
| Cases treated twice, 24 hours apart (Permethrin and BB) | 1 |
| Case treated once (Permethrin) | 1 |
| Cases treated once (Ivermectin) | 1 |
| **Treatment of close contacts** |  |
| No treatment available to anyone | 4 |
| All close contacts | 4 |
| Unknown | 3 |
| Symptomatic contacts | 1 |
| **Environmental decontamination** |  |
| Clothing | 7 |
| Bedding | 7 |
| None | 5 |
| Furniture | 1 |
| Shelters | 2 |

#### 3.2.1 Diagnosis

Eleven interviewees (92%) diagnosed scabies based on history and clinical findings. Two AHWs and the medical student confirmed their diagnosis with a doctor. One participant occasionally used skin-scrapings in a Greek formal camp and during an outbreak in Serbia. In one camp diagnosis was routinely carried out by a dermatologist and confirmed by microscopic examination. No participants reported use of dermatoscopy.

#### 3.2.2 Treatment

Three organisations attempted to follow Médecins Sans Frontiers’ (MSF) clinical guidelines for ordinary scabies treatment [27] (summarized in Appendix, p.5), one of which combined this with advice from the British National Formulary [28]. One organisation had their own scabies protocol. Only seven participants (58%) reported acaricide treatment was provided in camps they worked in whilst they were there. The remaining five (42%) reported only symptomatic management was offered (detailed in Table 1) because it was felt proper treatment would be futile due to high chance of re-infestation (four participants), or due to prescribing restrictions (one participant). In a third of cases treatment offered varied depending on availability. In NGOs that offered acaricide treatment, it was generally given to anyone complaining of itching, due to a low threshold of suspicion (5 of 7), however one NGO only treated individuals with a dermatologist-confirmed diagnosis, and one only treated ‘serious cases’.

#### 3.2.3 Outbreak Management

Eleven participants (92%) believed scabies was difficult to manage, whereas a GP following a formal camp’s scabies protocol considered it ‘just a nuisance’. Interviewees described high variation in outbreak management strategies (Table 2). This was particularly evident in timings used for permethrin application, which varied from singular treatment to second applications up to a week later. Three participants (25%) could not recall treatment timings or if close contacts were treated. In four interviews, participants claimed to treat all close contacts regardless of symptoms; any variation in treatment strategy was only mentioned once. Only three participants (25%) reported use of oral ivermectin for scabies treatment in camps, and this was limited due to lack of availability. Five interviewees (42%) reported environmental decontamination was considered too challenging, so was not carried out. Cleaning or replacing clothing and bedding was the most commonly used form of decontamination. Individuals were given replacement clothing/bedding and old items were washed (2 of 7), disposed of (3 of 7), or put into plastic bags for 72 h (2 of 7). One formal camp in Greece burned blankets. Another organisation managed to overcome the limited amount of clothing by building an inventory of rental clothes for individuals while theirs were in bags. Generally, attempted decontamination of furniture and shelters was not performed (only mentioned in two interviews). Where it was, it involved disinfectant or covering furniture and the insides of shelters in plastic sheeting for 72 h.

### 3.3 Camp characteristics and perceived barriers and facilitators to effective scabies outbreak management

#### 3.3.1 Camp Characteristics

All participants described camps as difficult places to work, and legal status seemed to have little connection to environmental quality. Participants described the poor standards of living experienced by camp residents (participant quotes, Appendix, p.6-8). For example, P8 described a formal camp (Greece):

> ‘It’s horrible. There’s only one water source for over two thousand people, there’s not enough clothes, people are sleeping outside, there’s not enough tents, the hygiene is terrible. I was there in the summer so it was hot, there’s not enough shading and people couldn’t find a place outside the sun. It’s more horrible now, as its winter, so it’s freezing, and there’s no heating. To be honest it’s worse than the refugee camps I’ve seen in Africa.’

P9, who had worked in both formal and informal camps (France and Greece) spoke similarly:

> ‘They are a human rights violation. I don’t think people should be living in them, especially in a first world European country this should not be happening. People are living in cold tents without heating and electricity, there’s rats, there’s scabies, there’s a lack of toileting facilities, a lack of hygiene, lack of food.’

Ten participants (83%) believed shelter was inadequate or non-existent, two (17%) thought it very basic. Twelve (100%) described lack of suitable sanitation and hygiene facilities. Five (42%) described worries regarding safety; either as a result of fights between residents, mistreatment from state authorities, or worry about stability of camp structures.

#### 3.3.2 Barriers and facilitators to effective scabies outbreak management

Themes and subthemes are illustrated with example quotes in Tables 3 (barriers) and 4 (facilitators), with further participant quotes in Appendix, p.9-25. Each interviewee described many barriers to effective scabies management. Four key themes arose; lack of Water, Sanitation and Hygiene (WaSH) facilities, healthcare barriers, social barriers, and organisational barriers.

**Table 3.**
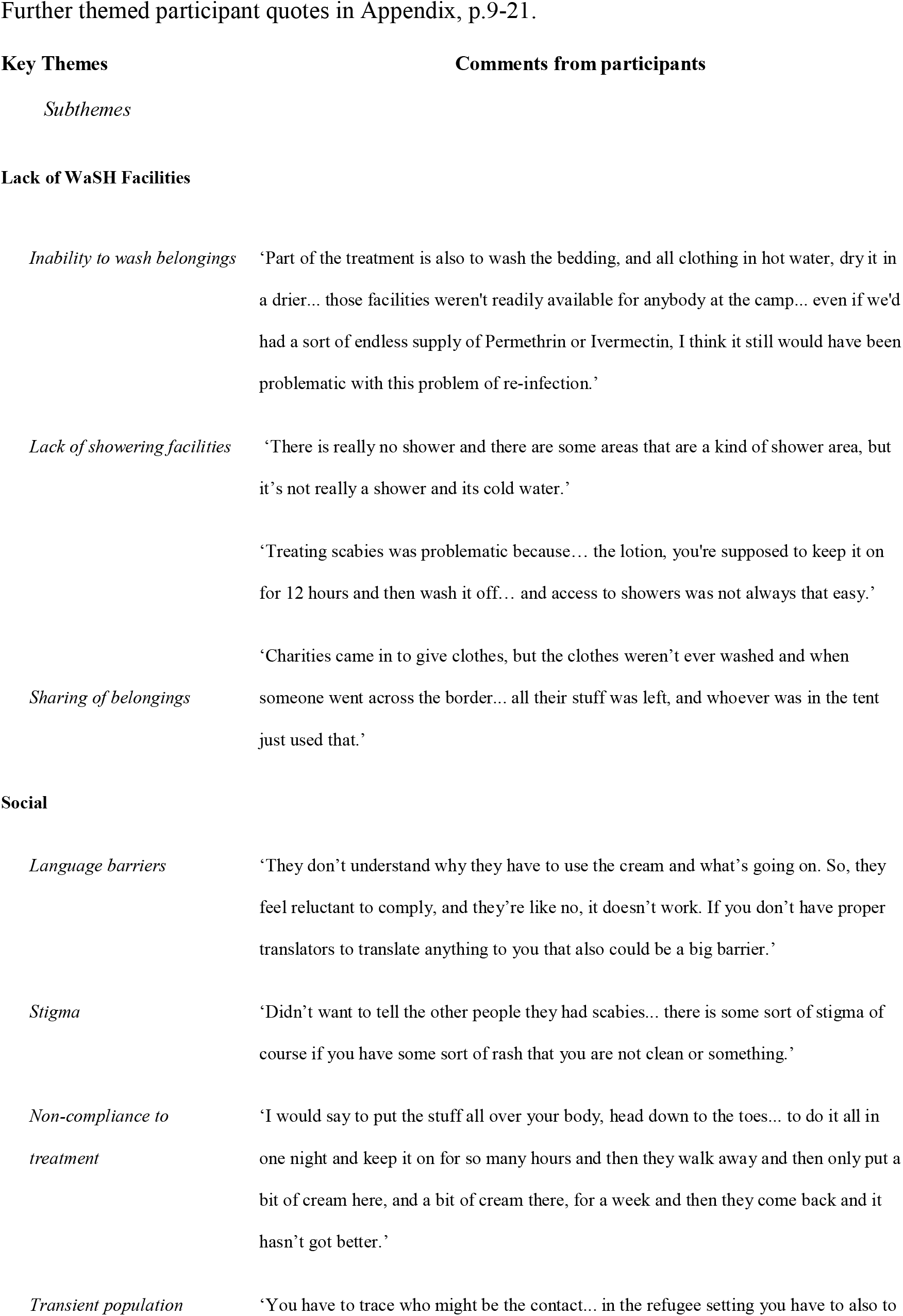
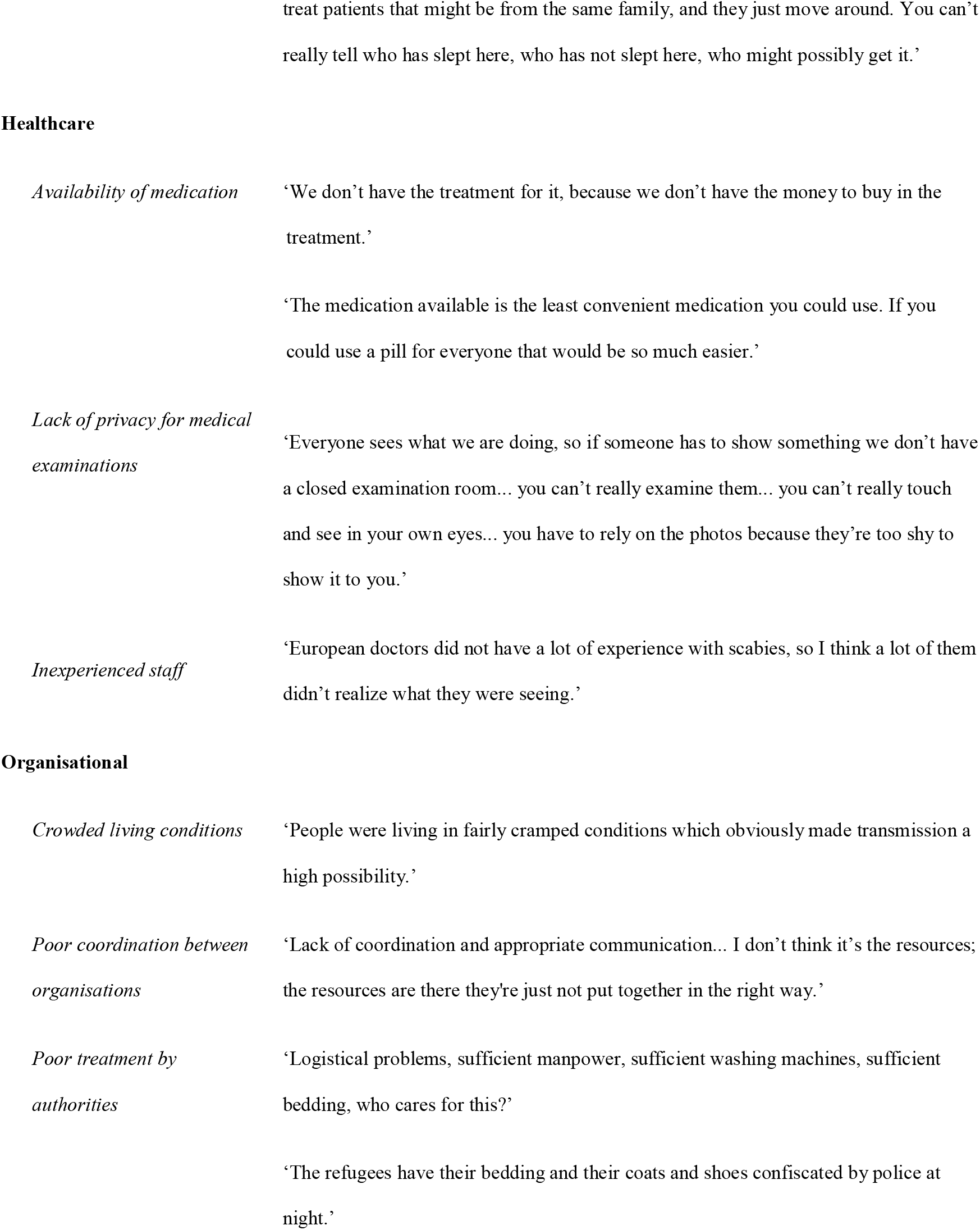
Barriers to providing effective scabies management

Lack of WaSH facilities: The main barrier identified by 10 interviewees (83%) was lack of access to washing facilitates for fabric belongings in order to prevent re-infection. Due to the lack of resources in camps, three interviewees (25%) described how sharing belongings was promoting re-infestation, which was amplified by the inability to wash them between owners. Six interviewees (50%) described the difficulties associated with inability for camp residents to wash themselves, due to non-existent or insufficient showering facilities. This was especially relevant regarding topical scabicides, which should be washed off [3].

Social barriers: Eight interviewees (67%) felt language differences were a major barrier, both in understanding individuals’ problems and subsequently explaining scabies treatment regimes and environmental decontamination. Four (33%) felt stigma associated with scabies infestation adversely affected management. Stigma reduced presentation to services and individuals were less likely to inform close contacts who required treatment. Treatment non-compliance was considered a barrier in four interviews (33%), as a result of complicated management regimes, and competing priorities for camp residents. One participant (8%) felt the solution was to have a protocol followed by all organisations, which allowed for continued community reinforcement of the chosen method within a camp. Mobility of the affected population was a concern for six interviewees (50%), who felt it affected contact tracing, follow-up treatment, and disrupted scabies elimination efforts.

Healthcare barriers: Eight interviewees (67%) considered scabicide availability to be a major barrier. Cost, prescribing restrictions, and variations in pharmacy stocking contributed to shortages, and for most NGOs availability relied on donations. Lack of consistency complicated treatment regimes and fostered mistrust of services. Three interviewees (25%) felt lack of privacy was a barrier to providing effective care. Some camp residents attempted to circumvent this by providing photos on phones, however this was not considered as useful as examinations. Five interviewees (42%) felt healthcare workers accustomed to working in Europe were not particularly familiar with scabies and therefore didn’t recognize or treat it correctly, and this was exacerbated by high volunteer-turnover.

Organisational barriers: Six participants (50%) perceived crowded living as a major barrier to effective management. Large dormitory-style rooms were perceived to be the hardest in which to determine close contacts, as were bigger units in which blankets had been used as partitions between families. Five interviewees (42%) described how poor coordination between organisations (both NGOs and governmental organizations) was a significant barrier to effective care. This primarily involved organizations providing different resources without communicating with each other about appropriate timings (e.g. lack of connection between timings of treatments, clothing replacement, and access to showers). Two participants described disrespectful care and stigmatisation by healthcare workers. Ten participants (83%) described how lack of support and mistreatment by state authorities acted as a major barrier, most of which was described in informal camp settings (7 of 10). Camp members faced police brutality (mentioned in two interviews), including having tents slashed, and clothing, shoes, and bedding confiscated. This encouraged sharing (with potential implications for transmission), and by adding further challenges to everyday-life it reduced the capacities of volunteers and patients to focus on treatment and decontamination regimes. It was believed local state authorities did not want to provide basic facilities necessary for effective scabies management as they didn’t want camps to stay. This went beyond medical neglect to actual obstruction of aid efforts in some informal camps. One participant reported authorities ‘sometimes they cut… water, they try not to get us to be able to give out clothes, they raided our distribution place.’ The participant’s explanation was: ‘What the government is trying to do is, they’re trying not to get the refugees too comfortable.’

Facilitators: Although two participants (17%) said they did not experience any factors that aided effective scabies treatment and management, the remaining described five key facilitators, when available; WaSH facilities, language facilitators, education, treatment incentives, and coordination of organisations (Table 4). Three (25%) felt access to showers and washing machines were vital to successful treatment and outbreak prevention. Limited resources, such as clothing and bedding could be reused without concern of reinfection, and topical treatment guidelines could be adhered to. Nine interviewees (75%) described the importance of translators in overcoming language barriers. Only three NGOs used official translators, the rest were aided by camp residents. One participant described using recorded voice messages and information sheets in different languages. Four (33%) believed education of residents played a positive role in successful scabies management, by alerting them to others who needed treatment and enabling them to educate them in correct treatment application. This awareness raising was also thought to have reduced associated stigma within the camps. Four (33%) stated incentives, such as receiving new clothing or just underwear, were helpful for treatment compliance, therefore minimising spread and re-infection of scabies. Eight interviewees (67%) reported the positive impact of communication between organisations, including information and resource sharing, and enabling direction of residents to appropriate organisations. The GP who described scabies as only a ‘nuisance’, attributed that to having a camp scabies protocol. In particular, the ‘ability to pass on a systematic and well-rehearsed message’ was thought to have improved adherence and general understanding, therefore improving effective scabies management.

**Table 4.** Facilitators to providing effective scabies management

Further themed participant quotes in Appendix, p.21-24.
|  |  |
| --- | --- |
| **Key Themes** | **Comments from participants** |

## 4.0 DISCUSSION

The crowded and substandard living conditions described by participants provide an ideal environment for scabies transmission, whilst obstructing treatment and outbreak management. Some participants believed lack of political will was preventing provision of suitable living conditions and access to healthcare as governments feared they would encourage people to stay too long. Going further, participants reported actual obstruction by authorities of camp resident self-care and NGO aid efforts, with implications for transmission and control of scabies. Without the support and coordination of all those responsible for the care of refugees/migrants, successful scabies management will remain highly challenging, and the resource poor and overcrowded aspects of camp environments which enable outbreaks will likely remain unchanged. Indeed, in late 2020 a situation similar to the ones described for 2014-2017 was observed by co-author CS on Lesvos, Greece, in the formal camp Kara Tepe 2. The NGO CADUS, in which CS works, had been deployed within a WHO initiative to provide medical care from 26.10-18.12.20. Scabies was widespread among the camp population of 7500, but the eradication program had to begin with a capacity of ten treatments per day. A small volunteer-staffed NGO provided the treatment, including provision of (camp-external) showers, clothes and blankets exchange. At the clinic itself, mostly only symptomatic treatment could be offered, which was perceived as highly unsatisfactory by CADUS medical staff. The overcrowded camp lacked adequate showers and facilities to wash clothes.

### 4.1 Diagnosis and treatment

No participants were aware of dermatoscopy use in camps which is not surprising given the expense of equipment, a lack of individuals with the necessary training, and reduced sensitivity of dermatoscopy when examination time is limited by outbreak size [29].

However, the 2018 publication of consensus criteria for the diagnosis of scabies provides a global standard for clinical diagnosis, though it will require validating in these settings [30]. Some participants suggested screening on entry to camps as part of routine medical checks, conducted in an attempt to reduce entry of new infestations [31]. This may be feasible in camps and has been carried out in reception centres specifically for scabies [32]. However, in informal camps screening may not be consented to by those seeking entry and NGOs may not have sufficient resources to adequately screen individuals, which may lead to false positives/negatives. In addition, the asymptomatic incubation period of scabies would mean some potential outbreak index cases would still be missed.

Participants reported that the use of topical scabicides caused problems for treatment and outbreak management in these settings. Firstly, topical scabicide treatment involves full body coverage with creams, left on for at least half a day. There is limited privacy and individuals living in close proximity may have wished to avoid being seen treating themselves for a highly stigmatised condition, and so have been discouraged in applying appropriately.

Secondly, topical scabicides need be washed off to reduce potential irritancy [3] but WaSH facilities were usually only minimally available (if at all). Mass Drug Administration (MDA) of oral ivermectin is easier to administer, and were conducted successfully in reception centres in the Netherlands and Germany during the same period [32,33]. However, ivermectin is expensive in Europe and unlicensed for scabies control in most European countries [11] despite a good safety profile [34,35], and inclusion in the 2019 WHO Model List of Essential Medicines [36]. The potential health impacts of chronic conditions secondary to scabies [6-8] also support the notion that timely MDAs in these settings will have the benefit of reducing long term health risks and probably overall health costs.

Participants reported widespread use of camp residents as informal translators, but this raises concerns about confidentiality, and ad-hoc interpreters often misinterpret or omit important medical information [37].

### 4.2 Outbreak management

Discrepancies in management reflect lack of widely accepted standard outbreak guidelines. Guidance [for example, 27,28] is often for individual households and does not make adaptions based on the logistical difficulties of camp environments. Guidelines for institutional scabies outbreaks exist, yet none are based on a systematic review of the published evidence [11]. This is also the case for the 2017 “European Guideline” [38]. More broadly there is a lack of research regarding the most effective interventions for preventing transmission of scabies to close contacts [39], particularly in refugee/migrant settings.

Organisations often favoured attempting to decontaminate clothing and bedding rather than shelters, yet living mites have been found in dust samples on floors and furniture [40]. Variance in reported decontamination reflects a sparse historic evidence-base [11]. However, new experimental work [41] could support uniform action. Where washing/drying machines are available (few camps in this study) ≥10min at ≥50°C will kill all *S.scabiei* mites and eggs, though detergents have little effect [41]. Lacking such resources, some of our participants reported isolating potential fomites with plastic bags/sheeting for 72 h, so as to kill the mites. Due to mite desiccation this may have been effective in temperate-dry settings, but if bags were left outdoors in the cold and damp mite survival can be longer. At the outer limit, in warm-humid conditions elimination can take >8 d [41]. Existing guidance should be updated. Popular narratives that posit a causal association between levels of personal body washing and becoming infested with *S.scabiei* are contradicted by evidence from epidemiology and experimental trials [42]. However, even though improving WaSH facilities is unlikely to reduce skin-to-skin transmission, it is nevertheless required to support treatment and control where transmission via fomites may be a risk, when topical scabicides are used, and to minimise secondary infections. It could also be expected to reduce transmission of other diseases.

### 4.3 Strengths and Limitations

One study limitation is that time constraints and recruitment methods may have prevented a fully representative participant selection. Firstly, because recruitment and interviews were carried out in English, excluding experiences from non-English speaking individuals. Secondly, because overall there was fairly limited number of participants, and though data saturation was reached we cannot be certain viewpoints are representative. However, as a qualitative study it succeeded in giving voice to the subjective perspectives and experiences of healthcare staff involved in this previously unstudied topic. One study strength was that the relatively long semi-structured interviews gave participants opportunity to articulate their perspectives relatively unconstrained by researcher presumptions over what may have been important to them about their experiences. Nevertheless, inference and public health recommendations based on this research must be pragmatic with limitations acknowledged. Telephone interviews prevented acknowledgement of non-verbal cues, but their use did enable international recruitment. The retrospective nature of the study may have introduced recall bias.

### 4.4 Future Recommendations

Our study demonstrates the importance of having clear protocols for scabies diagnosis, treatment, and management in refugee/migrant camps. Systematic evaluation of the evidence base must be combined with input from individuals and organisations involved on-the-ground to provide both high-quality and feasible guidelines that explicitly consider camp settings – both formal and informal. Given most participants were not doctors, guidelines need to be clear and understandable to everyone involved. Though topical treatments will need to continue to be provided for children and pregnant women, the difficulties associated with them in this setting could be avoided for most adult patients through wider licensing and availability of oral ivermectin. The 2019 ‘WHO Informal Consultation on a Framework for Scabies Control’ recommends populations with ≥10 prevalence should receive ivermectin Mass Drug Administrations [43], and we suggest this should include refugee/migrant camps. However, strategies for lower levels of endemicity in these settings should also be investigated. More research and advocacy for those living in refugee/migrant camps in Europe is required to help improve management of common health problems and quality of life. This is particularly relevant for managing scabies in camps where substandard conditions, inadequate resources, and lack of guidance can lead to poor quality of care and ineffective treatment. As indicated by our participants, much of this work is shouldered by small, volunteer staffed NGOs operating with minimal resources in highly challenging (often illegally occupied) settings. Without broad political changes, this is likely to remain the case, and we should reflect on how we in the wider healthcare community can better support these initiatives, and those they serve.

## Data Availability

Relevant qualitative data is included in the article and Appendix, along with summarised quantitative data. Machine readable study data will be available on publication at: https://sussex.figshare.com/

## AUTHORS CONTRIBUTIONS

This study was carried out as part of the work of the Scabies Research Team based at Brighton and Sussex Medical School, London School of Hygiene and Tropical Medicine, and University of Southampton. Authorship order is alphabetical by surname, except the first (primary researcher) and last (lead supervisor). For clarity we detail contributions using the CRediT Contributor Taxonomy (http://docs.casrai.org/CRediT), and provide employment and disciplinary descriptions. Conceptualization: JM, JAC. Formal analysis: NR, JM. Investigation: NR, JM. Methodology: JM. Project administration: NR, JM. Supervision: SL, JM. Writing – original draft: NR, JM. Visualization: NR, JM. Writing – review & editing: NR, JAC, MGH, SL, CS, SLW, JM. NR is a junior doctor; JAC a public health physician and clinical academic in epidemiology; MGH a senior research fellow [global health]; SL a research co-ordinator; CS is an aid worker; SLW a consultant dermatologist, and associate professor [infectious and tropical diseases]; JM a research fellow [public health; neglected tropical skin diseases].

## APPENDICES

Supplementary Material accompanying this paper includes: examples of news articles reporting scabies in refugee/migrant camps in Europe; facebook.com hosted HCP group recruitment targets; participant characteristics; Standards for Reporting Qualitative Research checklist; MSF clinical guidance for scabies; themed quotations from participant interviews.

## ACKNOWLEDGMENTS

We thank the interviewees for their time and openness with our study, and the administrators and members of the healthcare staff networks who advertised our study on-line. We are also grateful to our colleagues at Brighton and Sussex Medical School, Elizabeth Ford (for input into study design), Jessica Stockdale (for producing figure 2), and Maya Khan (for setting up our Facebook recruitment pages).

## FINANCIAL SUPPORT

This research was financially supported by Brighton and Sussex Medical School from internal funds and did not receive any specific grant from funding agencies in the public, commercial, or not-for-profit sectors.

## CONFLICT OF INTEREST

None

## Notes

### Competing Interest Statement

The authors have declared no competing interest.

### Author Declarations

Brighton and Sussex Medical School Research Governance and Ethics Committee approved the research (ER/BSMS3398/1).

