## Appendix for "Scabies outbreak management in refugee/migrant camps across Europe 2014-17: a retrospective qualitative interview study of healthcare staff experiences and perspectives"

Submitted as a preprint to medrxiv.org 4/2021

**APPENDIX**

Examples of news articles reporting scabies in refugee/migrant camps in Europe	2

Facebook.com hosted groups in which the study was advertised	2

Characteristics of researchers	2

Table D1: Standards for Reporting Qualitative Research (SRQR) checklist	3

Supplementary Table S3. Participants’ characteristics and settings	3

MSF clinical guidance for scabies	5

Themed quotations from participant interviews	6

**EXAMPLES OF NEWS ARTICLES REPORTING SCABIES IN REFUGEE/MIGRANT CAMPS IN EUROPE**

**Cooper C**. Calais crisis: Migrants are living in appalling conditions, say doctors: Scabies is spreading like wildfire. The Independent. 26 September 2015, London. [Accessed 10 Nov 2017]. Available from: https://www.independent.co.uk/news/world/europe/calais-crisis-migrants-are-living-in-appalling-conditions-say-doctors-a6668631.html

**Lichfield J**. French police clear hundreds of asylum-seekers from Calais migrant camps after outbreak of scabies. The Independent. 28 May 2014, London. [Accessed 3 Jan 2019]. Available from: https://www.independent.co.uk/news/world/europe/french-police-clear-hundreds-of-asylum-seekers-from-calais-migrant-camps-after-outbreak-of-scabies-9444407.html

**Newton J**. Migrants living in Paris refugee camp often used as a stop-off before heading to Calais and Britain are forced to move on by police over fears they were sparking a scabies epidemic. Mail Online. 1 June 2015, London. [Accessed 3 Jan 2019]. Available from: http://www.dailymail.co.uk/news/article-3105501/Migrants-living-Paris-refugee-camp-used-stop-heading-Calais-Britain-forced-police-fears-sparking-scabies-epidemic.html

**TABLE S.1** ***Facebook.com* HOSTED GROUPS IN WHICH THE STUDY WAS ADVERTISED**

| - Boat Refugee Foundation – UK Medical Recruitment - Care 4 Calais Volunteer Chat Group - CARE UK Charity (Refugee aid NW) - Humanitarians in the UK - Information Point for Lesvos Volunteers - Junior doctors – refugee medical volunteers - Medics for Refugees (former Medics for Greece) - People to People Solidarity – Dunkirk and small camps - RAISE – Refugee Action In Somerset East - Solidarity for Refugees |
| --- |

### CHARCTERISTICS OF RESEARCHERS

NR is UK junior doctor who had completed an MSc in Global Health prior to the study, along with training in interview skills and qualitative analysis. JM and SL (project supervisors) are Brighton and Sussex Medical School staff with prior research into institutional scabies outbreaks, as part of the Scabies Research Team led by JAC [9, 11, 29, 35, 36, 42]. JM has previously done voluntary NGO work with refugees, and lived in physically similar informal camps to those described in the study.

### TABLE S.2 STANDARDS FOR REPORTING QUALITATIVE RESEARCH [26] CHECKLIST

| No. | Topic | Page/line no(s) |
| --- | --- | --- |
|  | **Title and abstract** |  |
| S1  S2 | Title  Abstract | 1/5-8  2-3/32-59 |
|  | **Introduction** |  |
| S3  S4 | Problem formulation  Purpose of research question | 4-6/60-113  6/114-18 |
|  | **Methods** |  |
| S5  S6 | Qualitative approach and research paradigm  Researcher characteristics and reflexivity | 7-8/144-64  20/446-59; Appendix 2/53-60 |
| S7 | Context | 4-6/60-113; Figures 1 and 2; Appendix 2/30-47 |
| S8  S9  S10  S11  S12  S13  S14  S15 | Sampling strategy  Ethical issues pertaining to human subjects  Data collection methods  Data collection instruments and technologies  Units of study  Data processing  Data analysis  Techniques to enhance trustworthiness | 6/122-39; Appendix 2/Table S1  7/137-39, 8/165-69  7/140-55  7/140-46  8/173-80; Figure 2  7/155-59  7/150-55, 8/158-64  8/162-64, |
|  | **Results/findings** |  |
| S16  S17 | Synthesis and interpretation  Links to empirical data | 10-15/194-331  12/242-52; Tables 1-4; Appendix 9-25; |
|  | **Discussion** |  |
| S18  S19 | Integration with prior work, implications, transferability and contribution(s) to the field  Limitations | 15-18/333-407; 19-20/424-44  19/408-22 |
|  | **Other** |  |
| S20  S21 | Conflicts of interest  Funding | 21/479-80  21/474-77 |

**Table S.3 PARTICPANT CHARACTERISTICS AND SETTING**

| **Sex** |  |
| --- | --- |
| Female Male | 10  2 |
| **Age** |  |
| ≤30  31-40  41-50  51-60  ≥61  Unknown | 5  2  2  1  1  1 |
| **Type of HCP** |  |
| Doctor  Nurse  Allied health professional  Other | 4  4  3  1 |
| **Number of years practising as HCP** |  |
| 1  2  5.5  11  12  22  24  40  Unknown  N/A | 1  2  1  1  1  1  1  1  1  2 |
| **Nation state of usual practice** |  |
| UK  France  Belgium  Netherlands  India  USA | 6  1  1  2  1  1 |
| **Pay status** |  |
| Volunteer  Paid staff | 10  2 |
| **Legal Status of camp *** |  |
| Informal  Formal | 9  6 |

** n=12, total >12 as some participants worked in multiple camps*

**MSF CLINICAL GUIDANCE FOR SCABIES** [27]

All cases:

- Close contacts treated simultaneously
- Clothing and bedding changed after treatment

1^st^ line**:**

**Permethrin 5%** (lotion or cream)

Apply everywhere on the body, contact time of 8 hours then rinse

One application may be sufficient, second application 7 days later reduces treatment failure

Or	**Oral ivermectin** 200mg single dose

Single dose sufficient, second dose 7 days later reduces treatment failure

Not for pregnant women or children under 15kg

If 1^st^ line treatment unavailable:

**Benzyl Benzoate 25%** lotion

Diluted before application use based on age

Apply everywhere on the body, contact time of 24 hours then rinse (contact time reduced for infants/babies)

Second application reduces the risk of treatment failure (eg. after 24 hours, with a rinse between applications or two successive applications 10 minutes apart)

### THEMED QUOTATIONS FROM PARTICIPANT INTERVIEWS

Quotes from the thematic framework matrix, extracted from verbatim transcriptions of 12 interviews, 34-71min each (mean 47).

### Description of the Camp Environment

##### *General*

P1: “It was quite distressing to look at really, and people were exploited.”

“There was quite a few cameras from the newspapers... which was quite intrusive”

P4: “Can you imagine like frontline war correspondent, you know just getting there or I know frontline medic you know in World War 2, bit like that without the bullets”

P7: “Very confusing, very high stress levels and very difficult. It was a transient camp, people came straight from the boats, the tents, they were hypothermic, it was winter, so it was extremely transient and extremely a difficult situation.”

P8: “It’s horrible. There’s only one water source for over two thousand people, there’s not enough clothes, people are sleeping outside, there’s not enough tents, the hygiene is terrible. I mean I was there in the summer so it was hot, there’s not enough shading and people couldn’t find a place outside the sun. It’s more horrible now, as its winter, so it’s freezing, and there’s no heating. To be honest it’s worse than the refugee camps I’ve seen in Africa”

P9: “I think they are a human rights violation. I don’t think people should be

living in them, especially in a first world European country this should not be happening. People are living in cold tents without heating and electricity, there’s rats, there’s scabies, there’s a lack of toileting facilities, a lack of hygiene, lack of food.”

P10: “You don’t see why everyone says it’s horrible, it doesn’t seem that bad when you first arrive, but when you start talking to people who live in these camps every single day, you see how it crushes them over time.”

“The other thing that really struck me was how gender based vulnerability was accounted for in practice. When it snowed, they kept moving women out to a better and nicer camp, but men get cold to yet they only moved out the women. It sucks to be a man.”

P11: “It was like an emergency pavilion... they put 3,000 refugees together in a big camp... Every tent, there were 10 people sleeping, and they went eating in a large room, where they could eat together with 100s of people. They have all the place where they can go to the toilet, and places where they can wash themselves, but it was all outside, it was inside they could sleep and eat, but they can wash themselves outside, and they could to the toilet. It was all arranged, but it was very basic.”

##### *Shelter*

P1: “It’s very organised... you think you’re in a small village really... but then when you go beyond that you see just a sea of tents where people just live”

P2: “There’s not even proper tents or anything, they just use whatever they can find”

“In Serbia it’s almost like a jungle, so it depends on like how lucky you are. So, some refugees have tents, some doesn’t have tent. And then sometimes they just sleep in the field because they walk to the border and sometimes they just stop in between and they sleep like under the stars.”

P3: “There’s no shelter... there’s just literally people sleeping in bushes”

P4: “There are no more camps, they’re gone... so we have like, nine hundred refugees living rough in Calais and then we have probably about four hundred and fifty refugees living in the forests of Dunkirk.”

P5: “People live there with tents or without tents.”

P7: “The camp was steadily improving by the volunteer and NGO work, and residents work, it was not as stressful environment as the factory hole which had been utilised in the first as tents... It was not a pretty environment but a lot of effort to make it a habitual environment and make it was a better environment in the end.”

P12: “Different locations so it was the side of roads mainly, and next to the roundabout there would be a patch of grass and there would be a large group of people on the grass... then at another location might be in kind of a residential area but at the back of that there’d be a field, just some woodland and then there’d be a large group of people there as well so it was, it was groups of people scattered throughout Calais. It wasn’t a camp”

##### *Water and Sanitation*

P1: “Running water was in troughs, like animal troughs. People washed in the open, or didn’t wash at all. They didn’t have any clothes washing; there was no way... People would just wash their clothes and just hang it over a bush or something”

P2: “There is no proper latrine system”

“In Serbia, there’s one German NGO that are giving free showers every second day for refugees. They also provide some water so the refugees can wash their clothes and stuff. But, in general, in one setting they can only give showers to up to forty people max. So, there are like one hundred and fifty to two hundred people, they could only give showers to forty people. So, some people might not be able to do showers, like maybe you can do once in a week. The sanitation is quite bad, and even the water, sometimes there is no water, if this NGO doesn’t come and provide water, so they do take water from some cart, but it's not really clean”

P3: “There are portable toilets there, but that’s about it, really. Occasionally an organisation, like Calais government, kind of, a local authority, they occasionally bring, I guess you would describe them as, not showers, but more like taps, portable taps with cold water, so people could have a kind of a quick wash and things”

P6: “It was quite well established... having said that, facilities were still really quite basic. The toilets were just portaloos, there was very limited access to showers, and running water and it was very muddy”

P10: “I never stepped inside any of the facilities in the camps, I smelled them as I walked past but I didn’t have an incentive to go inside so I didn’t. I assume they were quite terrible, like I could smell them, but not good, I did not see them, they were not good, I didn’t want to see them.”

##### *Food*

P3: “There was the food distribution which was run by volunteers so there would be a van going twice a day to camps around Calais and once a day to Dunkirk. But apart from that, there’s very little”

##### *Safety*

P1: “There was areas there where the women and children stayed that was quite well organised and had security and were patrolled... but a lot of people didn’t want to go in there, and often families were separated”

P2: “Often the riot police, they’re called the CRS, they have these big vans and they would often kind of hover around, trying to intimidate the refugees and the volunteers, and so that didn’t kind of make for a very nice atmosphere it was quite tense”

P5: “It feels like the woman, she is not completely safe because they protect themselves, they are old, sometimes there are pictures of walls falling down. It’s not safe also because there are a lot of tensions so sometimes fights happen.”

P12: “People were hiding from the authorities... because tents were being routinely slashed and belongings were being destroyed by the police and authorities”

***Healthcare***

P2: “Those camps in Turkey, the settlements, they are really in bad shape, no NGOs are running in, no-one to give free food, no-one to give the healthcare. I mean we were there giving free healthcare, but basically we didn’t go and visit them every day, so they’d basically sometimes end up not having at all, and they were refused to get healthcare in normal hospitals”

P10: “You start to see how someone with heat failure isn’t able to access treatment - it’s crushing him. A person with diabetes can’t get their blood sugars up - its killing them.”

“A lot of organisations can be disrespectful, especially the doctors, they can be very humiliating, that makes people reluctant to go back for treatment. And they can also be rather indifferent to a lot of things, which means things don’t get taken care of. Refugees wait in line for several hours, to be seen, to be disrespected, so they don’t go back to the doctors.”

**Barriers**

#### Water, Sanitation and Hygiene Facilities

##### *Washing of clothes*

P1: “Clothes that they hadn’t been able to wash, or they only came over in their clothes that they were standing in”

“There wasn’t anywhere that people could wash their clothes, yeah, have washing facilities to wash their clothes... They wanted to be clean. And it’s very difficult to keep clean if you’re living outside in a tent”

P2: “Most of the scabies has gone so bad, the scar has got so infected because of the very poor clean and hygiene. They’ve been wearing the same pants, sometimes if it’s raining you get wet and you don’t have any new clothes so you just wear the wet clothes all over again.”

“They might have some utensils to cook, but not enough to boil so much of water, or somewhere they can soak all the clothes and to wash them. And even if they hand it all to the volunteers, even we don’t have the capacity to disinfect all the clothes”

“Sometimes they don’t have water...it’s quite tricky to wash the scabies infected clothes. If you have big washing machine that has a warm water setting, that would be great. But some of them doesn’t have it, so you have to boil water and soak it or you have to put chemicals and stuff... the chemicals that were needed, you don’t know where to get them, so you need to get support from the larger NGOs. If you don’t have larger NGOs then you might not be able to do it”

P3: “People have so little and they don’t want to give any clothes or bedding that they have up... definitely not clothes washing”

P4: “There was like cold taps there that they could wash with to wash it off the next day, but my problem with that was the clothes that they’re putting back on. Yeah, the clothes have to be washed at a high temperature. So it’s very much a struggle”

“the treatment we were giving was that because it was toxic we told them you have to wash it off the next day, that they could shower it off, but that’s all. They put back on infected clothes or they’d dry with infected towels”

P5: “Then we have other issues which is means we have to get clothes, new clothes for these people and new sleeping bags because, and then we cannot tell them to wash the sleeping bags in the residence at 60°, it doesn’t work because they’re homeless.... they can only wash their clothes by hand so of course they cannot wash everything that there is... we cannot provide clothes for all of them.”

P6: “Even if a patient is treated for scabies, part of the treatment is also to wash the bedding, and all clothing in hot water, dry it in a drier picking up all the scabies mites. And those facilities weren't readily available for anybody at the camp... I still think that even if we'd had a sort of endless supply of Permethrin or Ivermectin, I think it still would have been problematic with this problem of re-infection.”

P8: “The challenge was to do the hygiene part of the treatment... The problem was to find enough clothes for them to change their clothes because there is no washing thing in the camp so we would have to do the whole thing of putting the clothes in a garbage bag, close it for 72 hours, then take it out. Then we actually found out that a lot of people don’t actually have a spare set of clothes, or they have a sweater and trousers but don’t have more underwear.”

“There’s only one water source for over two thousand people... there was no washing facilities for the people, they had to wash their clothes by hand. There was no soap provided. People did not have the possibility to really wash their clothes well.”

P9: “They couldn’t wash and we didn’t have enough blankets to give out, so everything was getting re-infected all the time. We couldn’t take everybody’s clothes because there wasn’t enough clothes to give out again so that was really hard to manage... Things were just getting re-infected because it wasn’t very well controlled.”

P11: “Insufficient washing and bedding, and like not enough washing machines... a lot of people, most of them only had one pair of clothes, and they can't change them, when they treat. We need clean clothes, so that was a difficult part... You also have to think about the hats, scarves, they also have to wash them, because in the beginning, that was a thing we didn't clean the hats, scarves.”

P12: “Most quite overwhelming was the lack of amenities, people’s clothing, sanitation facilities, so no showers, no toilets.”

##### *Shower facilities*

P2: “There are one hundred and fifty to two hundred people, but they could only give showers to forty people. So, some people might not be able to do showers… the sanitation is quite bad, and even the water, sometimes there is no water. If this NGO doesn’t come and provide water, so they do take water from some cart, like not really clean and it’s not really in a good shape.”

P3: “Not having access to showers”

“That would be the only way to get rid of scabies. But this doctor said that he was already, I mean it’s a small clinic, he was already overwhelmed, he was saying he’d have to… They only had two showers, there’s no way they could do that, he just wouldn’t be able to cope with that many people.”

“They don’t have showers, or they don’t let the refugees use the showers.”

“At least in Calais they have these kind of portable taps, shower things. That’s not provided in Dunkirk, the council in Dunkirk won’t provide that for the refugees. One of the main barriers we faced was getting proper showers to people, or people to the showers.”

P4: “Lack of showers and lack of laundry meant that you couldn’t focus on one. Okay, here’s a tube of scabies treatment, it’s quite toxic so you put this on your body, rub it all over, twenty-four hours later you wash it off, but all your clothes have to be clean and they’ll look at you like really how. It’s a very, very uphill struggle to treat it and it’s really prolific.”

“They did have that facility [clothes washing] in the Dunkirk camp but, towards the end of the camp, before it burnt down, they stopped washing clothes and they stopped, they turned off the showers and then scabies started there as well... In the beginning in the Dunkirk camp it seemed manageable. When we were there we hardly saw any. It was under control, but then the minute you put people out there in the middle of a forest in filth and dirt.”

P5: “There is really no shower and there are some areas that are a kind of shower area, but it’s not really a shower and its cold water. Basically it’s not helping at all... it’s very, very difficult to stay clean.”

P6: “Treating scabies was problematic because as you're probably aware, with the Permethrin treatment, the lotion, you're supposed to keep it on for 12 hours and then wash it off after a 12-hour period, and access to showers was not always that easy... I never actually saw the shower facilities there but I understand they were very basic and there were often very long queues for the showers. So it wasn't easy to shower.”

P8: “There is not even one shower, there’s no soap to wash yourself... people couldn’t even wash their bodies with soap.”

##### *Sharing of belongings*

P1: “Charities came in to give clothes, but the clothes weren’t ever washed and when someone went across the border, tried to come into England, all their stuff was left, and whoever was in the tent just used that”

P2: “The main biggest problem is to get them to improve their own hygiene... to get them to clean themselves, and have each one have their own towel, and not to share them.”

P8: “A lot of people basically only had one pair or underwear, or socks, or they couldn’t wash their shoes... they usually only had one pair of blankets... There would always be something that wouldn’t be replaceable... Because of lack of things for themselves, they keep borrowing each other’s clothes and blankets.”

#### Healthcare factors

*Availability of medication*

P2: “Limited availability of medications.”

“In Turkey, we don’t have big NGOs because they’re not allowed to be… so that was a bit tricky. We need get all this medications on the camp by ourselves. It’s also depends on what is in each country. In some countries they use Benzyl Benzoate, then some of the countries Permethrin, so you need to know which one that they use for that country and what’s familiar to that country. Then you can source it locally... Some people smuggle it in the bag, and they took a flight and then they give it to the organisation. In the long run it’s not going to work because when you run out and you change to different regimes that’s a bit complicated, if you stick to one thing that’s easier to source.”

P3: “The medication was very expensive... there’s a lot of people, nearly 1000 people there in Calais and Dunkirk, that’s an issue.”

P4: “We don’t have the treatment for it, because we don’t have the money to buy in the treatment.”

P5: “Only one we can buy in Greece is the more difficult to apply and the one you have to apply many times.... so there is less chance of success of people really following how to do it.”

P8: “We didn’t have enough medicine most of the time... what we would do is write down their names and a week later if we got the medicine we would ask them to come back... Because we were short of pills we would just give it once.”

“In the beginning they trusted us, after a while they didn’t because we promised them treatment and we didn’t give it.”

P9: “Just didn’t have enough treatment... we have to fundraise to buy it, and if we don’t have funds we can buy it.”

P10: “Permethrin in pill form [we presume they mean ivermectin], they don’t sell that in Greece, that would be the most convenient way to provide medical treatment... BRF had a stock from Netherlands, but as soon as that ran out it was gone. The only treatment that is available is Benzyl peroxide, but you have to dilute that to make it the right concentration, and dilute it differently for children, then leave it on for 24 hours or at least overnight. There was only one pharmacy that would only sometimes have it in stock, so we had to arrange with them beforehand... The medication available is the least convenient medication you could use. If you could use a pill for everyone that would be so much easier.”

P12: “The lack of cream or treatment... Because none had been donate. We didn’t have any in the container, and we just weren’t able to get hold of any. The only medication we had was what was in the container, which I believe was from volunteers bringing over all donations. It was quite limited really.”

##### *Lack of privacy for medical exams*

P2: “Don’t have any proper building, clinics set up. So, everything sees what we are doing, so if someone has to show something we don’t have a closed examination room, we don’t have a private examination... you can’t really examine them, like you can’t really touch and see in your own eyes, you have to rely on the photos because they’re too shy to show it to you.”

P4: “You’re out there in the middle of, you know, you’re not somewhere quiet and private you can’t ask people to strip off.”

P12**:** “If you were treating somebody, there certainly wasn’t any privacy for people... you’d find there’d be sometimes, gathering around and watching you... amongst large crowd.”

“I don’t think we were openly saying to people, “Oh I think you’ve got scabies,” just because we didn’t want to say in large groups of people and I don’t think we wanted to publicise that type thing because you don’t want anybody to be offended or people, other people are listening.”

##### *Lack of staff with experience of scabies*

P2: “Most of the Western doctors are European, they’re not very used to seeing scabies in normal settings.”

“They have that scabies and it gets really bad and infected, some people might mistake it for like normal infected things... they don’t give the creams for scabies and stuff.”

“In Greece I met a lot of UK doctors. I find most of them, they know scabies and stuff. But when I was working with in Turkey and in Serbia, I worked with people from Europe, like Austrian and Germans, and then some of them had never seen scabies before. Because it’s not very common in these kind of areas... so in terms of that staffing, it’s not really hard to get staff, but it’s really hard to get staff that have seen or have managed scabies before.”

P4: “Most of the other first aid people they’d never seen scabies in their life.”

P8: “We heard they say they haven’t seen scabies which could mean they don’t know how to recognise scabies or they don’t want to, so could mean the other doctors in the camp don’t know how to recognise scabies.”

“Many people came to volunteer only for a few weeks so a lot of them didn’t have much experience to recognise it.”

P10: “European doctors did not have a lot of experience with scabies, so I think a lot of them didn’t realise what they were seeing.”

“A lot of volunteers had never seen scabies before... volunteer based organisations take people that want to volunteer, but because they need people they’re not really in a position to screen them for the job. I’ve definitely noticed that there really isn’t a lot of experience of doctors with scabies.”

P12: “I don’t know if I ever seen scabies before so I felt quite inexperienced.”

#### Social factors

##### *Language barrier*

P2: When I was working with the Afghan refugees, one of the time we couldn’t get a proper translator... and they don’t understand why they have to use the cream and what’s going on. So, they feel reluctant to comply, and they’re like no, it doesn’t work. If you don’t have proper translators to translate anything to you that also could be a big barrier to understand what’s going on really.”

“When I was working in Serbia we don’t have a proper translator, so we were using one of the refugees to translate for us, which is not very ethically correct I would say, because of the confidentiality issues”

P3: “There were a lot of people, their English wasn’t great and we rarely had someone on the team who could speak Arabic for example so sometimes it was difficult to diagnose someone... it was difficult for them to explain to us, and for us to understand exactly what the problem was.”

P4: “We didn’t have all translators... so that was an issue. We sometimes had to ask another refugees and call out for somebody that could come over and help and speak enough English to tell us what was wrong with this person... I couldn’t explain to him what I could do so it’d just be, just frustration.”

P6: “We didn't have proper translating facilities. Trying to explain all of what I've just explained to you about washing off the treatment, the lotion and washing bedding and clothing, even if that was all possible, explaining all that to people without proper translators was difficult.”

P8: “It’s hard, because we were a volunteer organisation we didn’t have official translators. We would always ask people in the camp to help us translate. Sometimes it was difficult to find translators in certain languages, like Kurdish. In the end that would mean that the Kurdish people have less access to our care than Arabic or Farsi speaking patients.”

P9: “We usually have translators... but you don’t always have the right translator.”

P11: “When you talk to 10 people at a time, and both women and men together, it's maybe difficult, and there are also people who had another language, and some languages are not known in other translator centre, so we couldn’t get a translation for them.”

P12: “Language barrier”

##### *Stigma*

P2: “To ask them to, not to share things with your friend, try not to sleep with them, I feel bad sometimes. They sleep together, they have like one blanket and all sleep like five people on the same blanket. So, when you say okay, you have this problem, you can’t sleep with your friend, it’s almost like giving them a stigma.”

P7: “Some people had social barriers, as in I am so clean and I don’t get this kind of thing... "I have eczema", or "I have dry skin" or this or that, "I don’t have scabies". Those were the people who often suffered for the longest... Possibly as a result of stigma surrounding scabies... Scabies and mites are not very pleasant things to have.”

P8: “didn’t want to tell the other people they had scabies... there is some sort of stigma of course if you have some sort of rash that you are not clean or something.”

P11: “it was difficult because the Syrian people looked at Eritrean people like they were dirty… because they had scabies.”

##### *Non-compliance with treatment*

P2: “This whole effort has to be put by the patient, and under these circumstances this patient has a lot … a lot other things to think about rather than this itchy thing ... The compliance is really hard, and also to get them to continue the treatment over a few days, also quite hard as well.”

P5: “You have to treat case by case explaining to them how to do the treatment with the message of giving them new clothes and sleeping bag and that are shown everything and even when we try to explain everything they don’t always do it correctly. That’s for a major issue because if the treatment is not done correctly then it won’t be effective.”

P7: “The one thing which we did encounter occasionally was people that didn’t listen to us, so I would say to put the stuff all over your body, head down to the toes and they got it everywhere... to do it all in one night and keep it on for so many hours and then they walk away and then only put a bit of cream here, and a bit of cream there, for a week and then they come back and it hasn’t got better.”

P8: “When they would hear about the whole treatment thing that their whole tent has to be treated or it doesn’t make sense, then they would say ok never mind and go away and don’t come back. Or people would come and say that their roommates don’t want to come, or they would bring their roommates and then the roommates would say oh this is too much work, I don’t have itching so I won’t take part in this.”

##### *Population Mobility*

P2: “You have to trace who might be the contact... in the refugee setting you have to also to treat patients that might be from the same family, and they just move around. You can’t really tell who has slept here, who has not slept here, who might possibly get it.”

P3: “Not being able to kind of, you know, everyone’s moving around Calais all the time, so not being able to kind of, I guess, not having the resources to be able to keep track of individuals with scabies.”

P5: “You have always new people coming in, and people leaving so it means that even the people treated, if they come to you and you treat them and then they disappear, not having any other treatment.”

“Making one day of treatment of scabies is not successful because the population keeps changing... all the energy and the money spent from the treatment day actually is a kind of a waste... in this population which is always changing, it is not a good use of resources because we won’t be able to properly medicate it.”

P7: “Often these [single young men] were the most transient people who would simply wander off at some point, and not come back for a second dose or not come back for collecting whatever they’re supposed to collect”

P8: “The fact that people move in the camp is also a big challenge because even if you treat a whole container, if one person is put there by the police and if they say this person has to sleep there and this person has scabies then the whole effort is for nothing. So that’s actually one of the biggest challenges, that people are moving all the time.”

P11: “It was difficult in this setting, because you can see there are 3,000 people, and everywhere probably 10 people in and 10 people out... it was also very difficult to treat them well, because normally we have to treat everyone together, at the same time.”

“It’s difficult to treat everybody together, and there's nowhere to isolate people, a lot of mobility of people, and logistical problems with that.”

#### Organisational Factors

*Living conditions* ***– dormitory style rooms***

P2: “In the government run camps they usually have their own tent or their own cubicle but in, in the jungle settings most of them live in quite overcrowded, they all just live, sit together and stuff.”

P3: “Barriers in terms of the crowded conditions... I think just in terms of people who are living close together so scabies is very easily spread.”

P6: “People were living in fairly cramped conditions which obviously made transmission a high possibility.”

P7: “The most difficult to treat was the single young men, of being in the large accommodation, and camp accommodation, while the families, were in the family rooms. The single young men were in large dormitory style rooms and so it was a lot more difficult to determine the close contacts.”

P8: “A lot of people were sharing tents or containers with large number of people.”

P10: “Sometimes you would have people living in bigger units... and people would partition of with blankets where different families lived. That was really difficult.”

##### *Poor coordination between organisations*

P5: “I remember one guy that they sent with everything for the scabies treatment and because it was my first day I was really trying to do it the way it should be done... He had an appointment for the day, so I wrote on a bit of paper and I wrote please allow this man to have two showers because he needs to do the lotion and wash it, 8 hours later. I sent him with a paper and everything. He had the new clothes, to bring back. Then they didn’t allow him to have the first shower because he had scabies. They sent him away because he had scabies, even though he was coming to get the treatment.”

P8: “A lot of things are managed by volunteers. Ideally in the camp you would want to have one named medical actor providing healthcare and asking for specialist to come and do clinics or to ask for advice from other organisations but there was no governmental healthcare facility. There was a huge gap because it didn’t even exist, so other organisations would come in and do little things but the whole coordination between us and the others were so strange... there’s no continuity. It was really unorganised.”

“So many different organisations with people coming and going, also the paid staff did not stay longer than 6 months to one year, and so many volunteers coming and going - there’s no continuity. It was really unorganised.”

P10: “Lack of coordination and appropriate communication. I don’t think it’s the resources, the resources are there they're just not put together in the right way.”

“Topical steroids was what was given for symptom relief because they couldn’t give anything else, proper treatment required too much coordination.”

“The medical organisation in Moria didn’t communicate with non-food item NGOs, so due to lack of coordination he could never have medication at a time that he wouldn’t be at risk of re-infection... X used to write prescriptions for non-food items that weren’t honoured by the non-food item NGOs.”

P11: “There was another NGO who worked there, and sometimes they did their own things, their own plan. Sometimes it didn't work well together.”

P12: “Clothing was distributed, but we weren’t really able to coordinate that because we, we didn’t really have communication between the individuals and the charity.”

##### *Lack of support and poor treatment from authorities*

P1: “There wasn’t anybody in charge of this camp, to tell people that there was a scabies.”

“They were hidden, you know, thousands of people hidden in one corner of France that one-one wanted, the whole world didn’t want to recognise.”

“Because the French Government didn’t want the camp there, they weren’t doing anything. There wasn’t even... basic facilities... They didn’t really want to provide healthcare for people. There wasn’t any shelter, there wasn’t any child protection, there wasn’t any protection for any vulnerable people there at all, adult protection, there wasn’t any of those type of things there.”

P2: [informal camps] “What the government is trying to do is, they’re trying not to get the refugees too comfortable, so sometimes they cut like water, they try not to get us to be able to give out clothes, they raided our distribution place.”

P3: “I mean a lot of people avoid going to anywhere where they’ll interaction with the authorities because, because of the police brutality, they’re worried about how they’re going to be treated and worried about being deported. Even though that can’t happen at hospitals and things, and people at this particular clinic, you know, they’re running it for people like this and they’re very nice, but there’s still that perception. I think a lot of people try to avoid going.”

“The refugees have their bedding and their coats and shoes confiscated by police at night.”

“We had to take someone to A&E once, and he was treated awfully by staff because he was a refugee... what we saw it was just the refugees being stigmatised at being refugees.”

P4: “In France you’re forbidden by law to give first aid, well not give first aid, but to give prescription medication, my hands were very tied into how to treat it.”

“Until authorities do the right thing and take care of these people properly you can never, will eradicate it. It will carry on as long as they can’t shower properly, as long as they can’t get clean clothes and wash the clothes the scabies won’t go away and the trouble is that the authorities in France and in Greece and even in Italy do not care and they’re not bothered and they don’t wanna know and basically refugees are depending on volunteer groups and small charities for their wellbeing and they’re very limited.”

P6: “Local authorities were not getting involved because the camp was informal.”

P8: “This official organisation that was responsible for that kind of camp, there was one doctor who was responsible for the public health in the camp, he was responsible but most the time he wasn’t there, he and the other organisation were ignoring that there was scabies in the camp... The Greek doctor kept saying it’s not a life-threatening disease, yes its annoying but there are worse things to care about, but even the worse things weren’t cared about... The people responsible for it would try to ignore it to ensure that they won’t have to deal with it... in the meantime there was lots of suffering.”

The care in the camp was very limited. In general, the government would be responsible to provide the healthcare but the government wasn’t there so they had contracted [healthcare] out... but they were always short of staff and short of a lot of things.

P9: “In the official camps if scabies happened it would be much easier to deal with because they have some washing facilities and they’ve got like WaSH and that kind of organisations working there. I was working more in non-official, and where it was happening more for me was nonofficial and we didn’t have that support.”

P10: “Lack of will and commitment.”

“A lot of organisations can be disrespectful, especially the doctors, they can be very humiliating, that makes people reluctant to go back for treatment. And they can also be rather indifferent to a lot of things, which means things don’t get taken care of. Refugees wait in line for several hours, to be seen, to be disrespected, so they don’t go back to the doctors.”

P11: “They were just logistical problems, sufficient manpower, sufficient washing machines, sufficient bedding, who cares for this? It certainly wasn't a priority for the organisation, because people have itching, but they are not going to die”

“The responsible authority, the central body for reception and asylum seekers... they don't support it.”

P12: “People were hiding from the authorities... the settlement or their place where they would sleep would not be there because tents were being like routinely slashed and belongings were being destroyed by the police and authorities.”

**“What would have made it easier to manage?”**

P1: **[**Washing facilities]

“I think it [washing facilities] would have made it easier to manage, because people could wash their clothes, and the people wanted to wash their clothes as well. They wanted to dry them, they wanted to wear clean clothes. They wanted to be clean. And it’s very difficult to keep clean if you’re living outside in a tent.”

P2: **[**Washing facilities; Public Awareness; Divided living areas]

“If you have a proper washing machine that have hot water then you can do it, or you can ask the refugees to do it themselves, but if you don’t have that then it’s really hard to be able to eradicate it.”

“It’s not hard, as long as you educate them and try to make them understand that what’s going on, this is what needs to be done.”

“For public health education you’d probably want to print out like a really easy version for it, or maybe you have to make a new one so they can hand out to the refugees to say we have these issues, and this is what happens if you are itching.”

“If you have like a proper segregations of rooms, or cubicles for each family, that will be a really good point to really manage it. You can just do entire families. It’s easier to also plot where the scabies is, you can trace them, where they go and what the patterns... But in the jungle they just get it like a wildfire.”

P3: [Coordination of organisations; Access to resources]

“If we could kind of put a system in place, if we worked with X, one of the main organisations at Calais, they have a warehouse and do clothes distribution... to see if we could organise, maybe have a ticket system so organise getting people treated at the clinic so with cream or tablets, and then on the same day giving them a shower, clean clothes, taking away the old clothes to burn... giving them clean clothes, new clothes straight away and giving them a ticket so they can come back the next day for another shower ‘cause you need to, and then to completely get rid of it, and making sure everyone in the camp does that.

“Access to resources... showers, clean clothes, and cream.”

P4**: [**Better living conditions; Washing facilities]

“If they had somewhere decent to live... If they had somewhere with a roof over their heads and if they had somewhere where they could have a hot shower and somewhere to wash their clothes.”

“I did think at one point I wonder if I could get like a mobile clothes washing van or something, it crossed my mind that it could be one thing we could have done with the treatment - get the clothes washed, shower, clothes washed, but it didn’t happen.”

P5: [Screening on arrival; Washing facilities]

“When we have treatment and sleeping bags and clothes, it’s easier to manage because at least we can do something.”

“The only measure that you could take is to take every new arrival and any sign of scabies and treat them, but it would take a lot of time to get sorted to do it. It is what they are doing in Serbia where all new arrivals with scabies, is quite a lot. We give all our new clothes and sleeping bag.”

“If we could have a place to send people where there is a washing machine and dryer and maybe a shower, in this case we could treat more because we could have, like the new clothes but if all clothes that people wear, and the sleeping bags and blankets.”

P6: **[**Oral medication]

“Availability of Ivermectin may have, to some extent because it doesn't require washing the lotion off afterwards, you know it's an oral treatment, a single dose oral treatment, may have helped but it wouldn't have really have overcome the problem of reinfection.”

P7: “Nothing in particular - it wasn’t a problem.”

P8: **[**Screening on entry; Cooperation of organisations]

“In every camp where people arrive there’s always a medical check for all the people so that would be a great time to check everybody for it, before they allocate them in a tent.”

“Cooperation of the people that manage the camp because they could help by not moving people all the time, or always discuss it with a public health responsible person in the camp, but there was no such person. Making sure to allocate people in the right place to not disturb the scabies treatment, that would be great. Working together, putting together resources, but the first thing is to recognise that there is a problem.”

P9: **[**Better living conditions]

“They shouldn’t be living in these conditions in the first place. They should be in homes and houses, not refugee camps and abandoned buildings.”

P10: **[**Interpersonal communication; Coordination of organisations]

“I think it’s really important to train humanitarian workers in communication and interpersonal interaction... You have to really care, you have to understand that the other person isn’t always going to understand right away and you have to work with them... There’s a lot of conversations that need to be had.”

“You need to appreciate what others in your NGO and what other NGOs are doing, and be conscious of the fact that it is a collaborative goal.”

P11: **[**More resources; Systematic treatment]

“If we had the time, the people, and just one tent more, then it would be no problem anymore because you had to do one day, very intensive, and a good job, and then you have not left problems... The plan I made I think it would be good, if there was money for it, if there were the people for it, and if there were an extra tent over there, then it was managed in one day, in two days. Then we would have to keep doing it, that every person who came in, and had the same treatment. You had first to do it with all the people who were there, Eritrean people, and after that you had to do every Eritrean who came in, we had to give them treatment, and then the problem was solved.”

P12: **[**Coordination of organisations]

“We could organise what the warehouse felt getting them fresh clothes, but then we’d have to also organise things like showers.”

**Facilitators**

P3, P4, and P12: “nothing”

***Water, Sanitation and Hygiene***

P5: “What was really helpful is that the centre where people could go with a shower, and have a washing machine... they can get a hot shower two times a week and they can also wash their clothes over there. I mean if we provide one set of new clothes, we can help them to wash the clothes they have. Twice a week is not enough to really make the treatment as it should be, but it is very good, because before we had only places with appointments for the shower, there was only one shower. There we can be sure they have their follow up of 10 days so they can get the treatment, the new clothes, the new sleeping bag and take to the shower.”

P7: “People have access to showers so that was not a problem, clothes had to be washed in the washing machines… and bed sheets and everything else was taken out and burnt. That was the protocol and we didn’t have a persistent problem, we didn’t have outbreaks.”

P9: “There was mobile showers we were getting access to, not everybody could get access all the time, so that was still difficult but there was that there so it’s better than nothing.”

#### *Coordination of organisations*

P1: “What we suggest we do is that they have clean clothing ready so that when they go to the Government organisations, health organisations for the scabies, that they can actually start treatment that evening, and we explain how they do that.”

“We referred a few people to A&E because they were quite unwell. We gave letters to people about scabies to bring to the medical place the next day. We made sure everyone had a letter saying it was scabies, and documented when it was and how many people were in their tent etc.”

P2: “We worked a team that provide showers. On one of the days, we call it a scabies shower, so where we have all the scabies patients. We give them a card that says if you have scabies you get showered tomorrow. We give them a card that says they have scabies, we give them a cream 24 hours before and then asked them to get showers, and then we put the creams again. So we partner with shower teams, and also, we partner with people from the distribution team so we could get them new fresh clothes. I think the treatment of scabies, it has to be like not just a healthcare provider, and it has to be like non-NGOs to help you work on that as well.”

“It’s really good to liaise with the large NGOs, that could provide, most of the time they [medications] were available. They have all those creams.”

P3: “NGOs were more of a help, as I said X said that they could provide clean clothes and they could work with us.”

P5: “Make a letter for them saying that this person has been diagnosed with scabies, please wash his clothes, allow him to have a shower and we will make a thing that it’s not as contagious and that you cannot catch it like this. It’s working well.”

P7: “There was the camp administration which was run by a local NGO and the staff were aware of the problem and were good at directing people to us if there was a problem... we gave the patients who were referred slips to take to the administration to get new blankets, new bedding and that and take the old stuff away for washing or burning and give access to machines for to the washing.”

“They have no difficulties staying on top of it, because the camp administration are behind it and the residents are behind it... That was all positive... the support from the camp administration.”

P8: “The willingness of other organisations like the warehouse on lesbos that provided us with clothes.”

P10: “Rented them clothes... built up an inventory of clothes, I worked with contacts in the warehouses to get donations so we could ship them over so we could lend them clothes based on their size/gender etc.”

P11: “We had a meeting with them [NGO]... they had their own plan, and it was different from our plan. But we talked about it, and then it's okay. We gave everyone a clear task and then the problem was solved.”

#### *Education*

P1: “We explain how they do that [the treatment] … and then for those that have been infected, for them not to shake hands, not to share bedding.”

P2: “If you educate the patient very well, make them understand very well what’s going on, and what needs to be done, I think that also worked very well... They understand, and they start to even manage to recognise their friends who have the same thing, which is really useful.”

P3: “Friends might have been able to tell each other. I don’t know help diagnose each other or encourage them to go to the clinics.”

P7: “One treatment which was once easily explained and where the neighbours could explain to you, and everyone could explain to each other and reinforce the message which we gave every day.”

#### *Language facilitators*

##### Translators

P1: “We had lots of translators there. We had our own translator who spoke several languages, and there were people who lived in the camp, working or volunteering as translators, and they were actually very good. Certainly, got the message across. So, there wasn’t a barrier.”

P3: “Some people did speak very good English and often if someone didn’t, they would get someone who they knew did speak English and they would come and translate, so that helped.”

P6: **“**Sometimes there were people that helped with translation, other occupants of the camp, other migrants would help with translating, but that wasn't consistent help it was just if they were around at the time and they offered to stick around to help a little bit.... if we did have people to translate then we were able to explain what the probable cause was of the itching and I suppose in itself having an explanation can offer some relief to people, and also reassurance that there wasn't anything serious in terms of, you know life threatening.”

P7: **“**They had very good translators for the other doctors and nurses.”

P8: “A lot of people came to help us translate... The few people who spoke English would always be after other people to translate. Many people don’t have anything to do all day so they’ll help to translate for other people.”

P9: **“**There’s a range of languages, if somebody knows both they can tell us a bit more.”

P10: **“**We were very lucky because X worked with unofficial translators from every community in the camp, so they were a great facilitator for getting the message across.”

P11: “The language is difficult, but we talked with the translator, with a live translator.”

P12: “There were a number of people there who had been residents in the UK but then had been deported so often there was somebody who could speak really, really good English. They would act as a translator and that was really helpful.”

##### Voice messages

P5: “Voice messages in different languages.”

“I tried to treat and tried to improve the way we are treating, to make it clear for the people in their language. So we have voice messages explaining how to take their cortisone treatment and how to apply the treatment. Basically for each person that we treat, we say in the voice message and ask which one [language] they prefer and then we play the voice message.”

##### Information sheets in different languages

P5: “We do have information sheets... we show them with the data, and the map to go to the shower place and there if this is written in Urdu, then we will have some positive treatment.”

#### *Incentive to take part in treatment*

P2: “The best way that we come up with is just get them to get rid of whatever clothing and bedding, and then we give them replacements back. So there’s an exchange there. That’s the only way that we can minimise the spread and stuff.”

P7: “The patients knew that it could be treated, some used it possibly as a way of getting new things and were quite disappointed if they didn’t have it, but that didn’t happen that often either.”

P8: “Some people when they found out we would give them another set of clothes and we would buy underwear for them, some more people were interested as they had an incentive to take part in it.”

P10: “We would take back everything except the underwear, so they got to keep the underwear, which for a lot of people was like an incentive to participate.”

#### *General*

#### Division of living areas

P2: “If you go for like a government run camp you have proper dividing, like in Northern Greece, they have cubicles. So, usually the outbreaks are a lot smaller and usually limited to a few cubicles, or maybe just one cubicle.”

“If you have like a proper segregations of rooms, or cubicles for each family, that will be a really good point to really manage it. You can just do entire families. It’s easier to also plot where the scabies is, you can trace them, where they go and what the patterns... But in the jungle they just get it like a wildfire.”

P7: “People were living in individual, family individual cells, not exactly houses, not exactly rooms… Anyway there was some level of physical insulation from family to family and so you could say this family is all close contacts and it gives you good conscience to clear other people like neighbours, declare them as neighbours, but not close contacts.”

##### Technology

P2: “We don’t have a closed examination room... But some refugees are quite smart, so they bring in photos.”

##### Donations

P3: “We managed to raise money for the cream... we have managed to raise quite a lot, well not a lot, but some money to be able to buy medication.”

“There would be donations of clothes, quite often from the main two warehouses.”

P5: “When I got the donation of ivermectin coming, it was good. I was so happy, I think the person who was leaving me so meds, she didn’t know what happened, I was really quite… I knew that it was so much easier to treat scabies.”

P8: “Our organisation runs on donations so if we did have a donation then we would be able to buy medicines.”

P9: “Fundraising for medicines.”

P10: “We had a stock [medication] from the Netherlands from donations.”

#### Guidance

P**7**: [Treatment protocol]

“The protocols worked well... The ability to pass on a systematic and well-rehearsed message.”

“One treatment which was once easily explained and where the neighbours could explain to you, and everyone could explain to each other and reinforce the message which we gave every day... We had one treatment which fits into the medical centre and one treatment which was approved by the different governments and that treatment is positive.”

P10: [staff advice/screening documents]

“Screening questionnaire that BRF staff could use to screen people, which was really important as a lot of volunteers had never seen scabies before... it meant that a lot of people could do the counselling, and tell people to get seen and at the time when the medication was being distributed the one or two doctors who had experienced seeing it would be there, but in the meantime it gave everyone else a tool to find household who had members suffering from scabies.”
